# Overexpression of miR-3168 impairs angiogenesis in Pulmonary Arterial Hypertension: Insights from circulating miRNA analysis

**DOI:** 10.1101/2024.04.30.24306656

**Authors:** Mauro Lago-Docampo, Ainhoa Iglesias-López, Carlos Vilariño, Adolfo Baloira, Joan Albert Barberá, Isabel Blanco, Diana Valverde

## Abstract

**Background:** Pulmonary Arterial Hypertension (PAH) is a rare disease where the thickening of the precapillary pulmonary arteries ends up inducing right heart failure. Nowadays, obtaining an early diagnosis is challenging and typically delayed until undergoing right-heart catheterization.

**Methods:** We performed small RNA sequencing (microRNA-seq) in the plasma of idiopathic PAH patients and controls, that we validated by qPCR. We then interrogated the role of miR-3168 in HUVECs by performing western-blot, flow cytometry and tube formation assays.

**Results:** We found 29 differentially expressed microRNAs and validate 7 of them let-(7a-5p, let-7b-5p, let-7c-5p, let-7f-5p, miR-9-5p, miR-31-5p, miR-3168) in a nationwide cohort of 120 patients and 110 controls. We then used classification models to analyze their potential as PAH predictor. In the first half of our cohort, we obtained a model with an AUC of 0.888. Although, this value lowered to 0.738 after using this model in the whole cohort of patients. Additionally, we validated the effect of miR-3168, a novel upregulated miRNA in PAH patients which targets *BMPR2,* and impairs angiogenesis, as assessed by the tube formation assay.

**Conclusion:** We identified novel downregulated and upregulated microRNAs in idiopathic PAH patients, developed a 3-microRNA signature for diagnosis, and validated *in vitro* that miR-3168 targets *BMPR2,* thereby impairing angiogenesis.

## Background

PAH is a rare and devastating disease characterized by the thickening of the precapillary pulmonary arteries. This induces an increase in pulmonary vascular resistance that eventually results in right heart failure [1]. The molecular basis of PAH is complex, genetics play a big role on it as the heritable (HPAH) and some idiopathic (IPAH) cases can be explained in a high percentage by mutations in an ever-expanding set of genes [2].

PAH early symptoms include shortness of breath, fatigue, weakness, angina and syncope, which are common for a wide array of disorders [3]. This complicates early diagnosis, often leading to delays as confirmation requires right heart catheterization (RHC) [4]. An early diagnosis and treatment is crucial to improve survival and quality of life during disease progression [5].

Several initiatives have been undertaken to enhance the diagnosis and classification of PAH using various-omic tools in plasma and serum. However, the only biomarker currently recommended for predicting prognosis in PAH is NT-proBNP [3], which is a marker of heart failure that correlates well with right heart dysfunction [6]. The quest for novel diagnostic markers began with the study of the metabolome to phenotype IPAH/HPAH against healthy controls. This approach successfully identified differences between the groups and also predicted the risk of early mortality and response to treatment [7]. Similarly, proteomic profiling aided in the classification of different PAH phenotypes based on variations in immune proteins [8], or even for early diagnosis of PAH associated with systemic sclerosis [9]. More recently, RNA sequencing in whole blood also demonstrated its potential in distinguishing PAH and predicting prognosis [10].

MiRNA are small non-coding RNAs of around 20-22 bp that regulate gene expression through binding to the 3’untranslated regions of genes [11]. They can be found in plasma circulating bound to Ago2 proteins, inside apoptotic bodies, and vesicles of different types. The diagnostic potential of circulating miRNAs has been proposed for years across a wide variety of diseases. PAH is no exception to this trend. In the initial approach, a miRNA array was used to compare lung tissues from individuals with corrected septal defects who did not develop PAH to those who did. This analysis identified miR-19a as a potential marker for PAH [12]. Not much later, researchers worked with an array to test miRNA levels in plasma samples taken right after the RHC. They found that miR-663 was a good classifier of response to O^2^ and iNO challenges [13]. In a different way, circulating miRNA have also been tested in the right ventricle and the pulmonary circulation, finding differences depending on where the samples were taken [14]. However, the latest advancement has been the development of a miRNA signature for PAH diagnosis using treatment-naïve patients. This approach ultimately identified 2 novel miRNAs (miR-187 and miR-636) with a clear association with PAH [15].

In this article, we utilized microRNA-seq to examine the differences in circulating miRNA levels between idiopathic PAH patients in functional classes-NYHA I and II and healthy controls. Our objectives were to identify the hallmarks of these early functional classes and discover novel miRNAs implicated in the disease.

## Methods

### Description of the cohort

For the RNA-seq, we selected 10 patients from the Vigo area and 15 from Spanish PAH registry (REHAP) biobank (hosted at IDIBAPS), all in functional classes-NYHA I and II. The 10 healthy controls matched in age and sex our patients; the controls were kindly supplied by LM-R from the Instituto de Investigación Sanitaria de Santiago de Compostela.

For qPCR validation, all 110 patient samples were from IDIBAPS and the 110 controls were supplied by the Spanish biobank network, samples were matched in age and sex. The study was conducted after the approval of the *Comité de ética da Investigación de Galicia* (CEIC Galicia) in accordance the clinical-ethical practices of the Spanish Government and the Helsinki Declaration.

An overview of the study design is depicted in Figure 1A.

**Figure 1.**
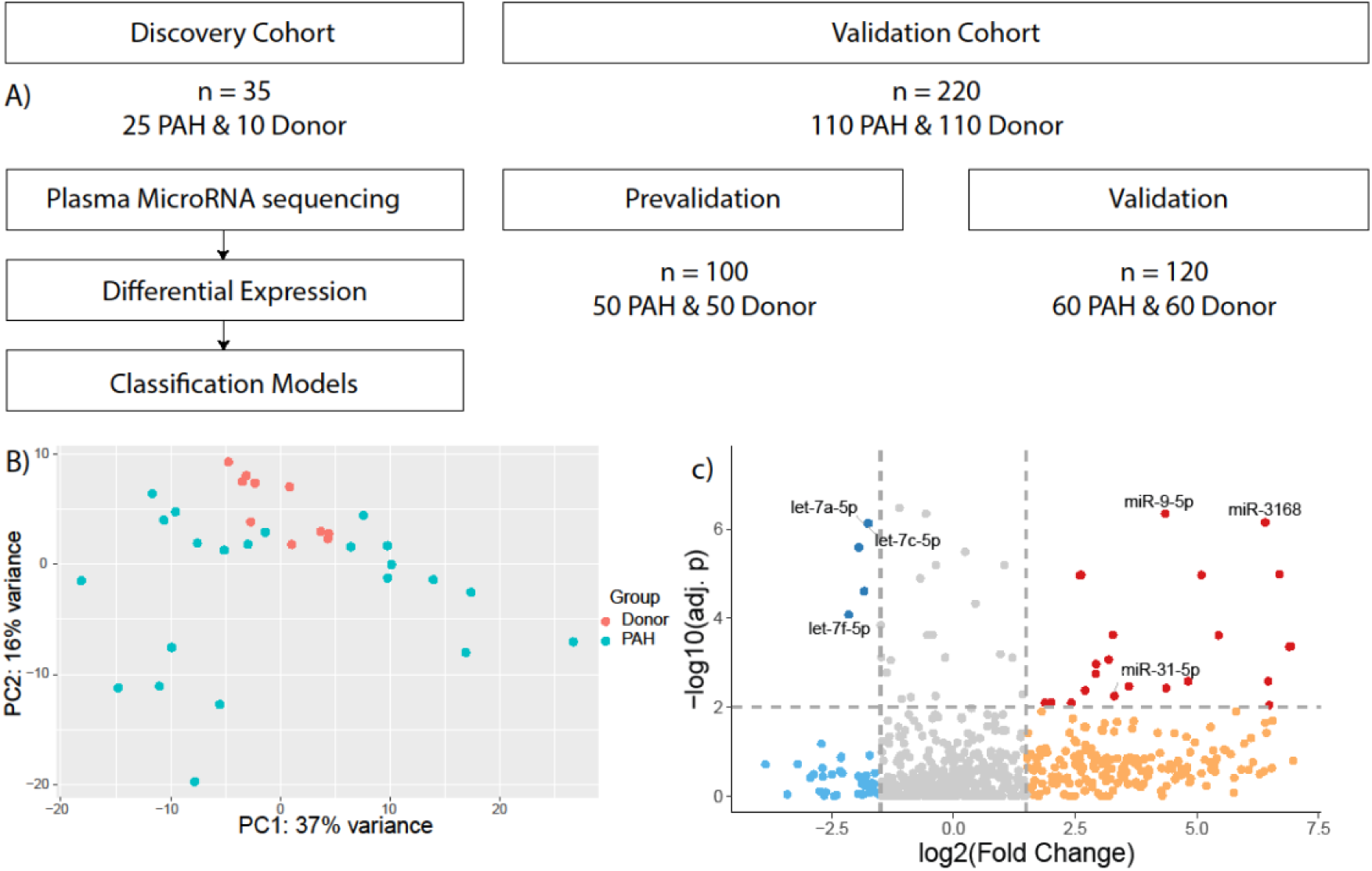
A) Schematic representation of the study. B) PCA plot of the microRNA-seq comparison between donors and PAH patients. C) Volcano plot representing differentially expressed miRNAs between donor and PAH. The labelled miRNAs underwent validation by qPCR in the REHAP cohort. X axis shows Log2(FoldChange), Y axis shows −Log10(adjusted p value). Grey represents the miRNAs that did not reach statistical significance (adj. p value > 0.01) and FC betwen ±1.5, Dark blue represent miRNAs that are significantly downregulated (adj. p value > 0.01 and FC −1.5), Red represents miRNAs significantly upregulated (adj. p value > 0.01 and FC 1.5).

### RNA extraction and microRNA-Sequencing

We centrifuged the plasma samples at 15.000 x g for 15 min to get rid of cellular debris. Then, we isolated 450 µl in a new tube and proceeded to microRNA extraction using the miRNeasy Advanced Serum/Plasma kit (Qiagen) following manufacturer’s protocol. After extraction, we quantified the miRNAs using the Qubit MicroRNA assay (ThermoFisher) and sent 10 ng for library preparation.

Libraries were prepared by RealSeq Biosciences (California, USA) using an in-house protocol of an Illumina Small RNA-like kit. Quality control was performed for all the libraries before sequencing, a total of 10 million single-end reads were expected per sample. Samples were analyzed in an Illumina - NextSeq 500 v2 High-Output - SR 75 Cycle (Illumina).

Reads were trimmed using CutAdapt 2.3 and aligned to miRBase with bowtie[16,17]. Then, we used Samtools 1.7 to convert SAM files to BAM, we built an index using Picard to convert the BAMs to read files, and used FeatureCounts (from the RSubread package) to obtain the count matrix for each miRNA[18]. Finally, we carried out the differential expression analysis using DESeq2 and EdgeR following the recommended protocol with a likelihood-ratio test to obtain the adjusted p-values[19,20]. The results between the two methods were compared and those miRNAs common between them were considered for the next steps. All the plots were generated all using the ggplot2 package in R [21].

### cDNA synthesis and qPCR

After elution of the miRNAs, we used 2 µl of the freshly extracted miRNAs to obtain cDNA using the Applied Biosystems TaqMan Advanced miRNA cDNA Synthesis Kit (ThermoFisher) following manufacturer’s protocol. The kit adds a poly-A tail to all the miRNAs, ligates an adapter and performs a RT. After obtaining the cDNA, we did a preamplification of all the miRNA+adaptor complexes using the Taq and universal primers provided by the kit. Finally, we diluted 1:10 the PCR products in nuclease free water.

Data were analyzed using the 2^-ΔΔCt^ using miR-103a and miR-16a as reference miRNAs. These reference miRNAs were selected using the data from the RNA-seq, looking for high count numbers and low FC difference between patients and controls. We then applied a log_2_ transformation to the normalized values before applying the classification models. To plot the individual miRNA levels, we did a Fold Change (FC) comparing patients against controls.

### Classification models and statistics

All the statistical analyses were performed in R (version 3.6). During the prevalidation, we performed a logistic regression using the miRNAs amplified by qPCR. Then, we used a Lasso regression to limit the number of variables in the predictive model. Based on the coefficients estimated in the Lasso, we generated models reducing the miRNA variables one at a time. The formulas to calculate the risk of PAH were built with these coefficients. To estimate the predictive ability of the models we carried out a ROC analysis using 1000 bootstrap replicates with the “pROC” R package to obtain the AUC [22]. In the validation stage, we tested the model with 50 different patients and controls. Then, we reformulated the model using 70% of all samples, leaving 30% to analyze the ROC curves and estimate sensitivity/specificity.

### Correlations and calculation of Risk according to the COMPERA study

Using the clinical data from the prevalidation and validation cohorts, we calculated the Risk following the criteria described in the COMPERA study[23]. Briefly, we used the NYHA functional class at diagnosis, 6-minute walking distance, NT-proBNP, BNP, cardiac index, and right atrial pressure. We assigned the levels of risk according to the cut-offs and used the R package “corrplot” (https://github.com/taiyun/corrplot) to visualize the correlations.

### Target prediction and functional assays

We used miRBase to predict the targets for the dysregulated miRNAs we detected [24]. Based on the lack of information we decided to analyze miR-3168 due to its predicted binding to *BMPR2*.

### qPCR and Western-Blot

To be certain of the results obtained in the luciferase assay, we transfected HeLa cells with 20 µM/40 µM of miR-3168 mimic (#HMI1108, Sigma-Aldrich, USA) or the scrambled miR (#MCH00000, Applied Biological Materials, Canada) using Lipofectamine RNAiMax (ThermoFisher). After 48 hours, we extracted mRNA with the NZY total mRNA isolation Kit (NZYtech) and performed a RT-PCR using the NZY M-MuLV First-Strand cDNA synthesis kit (NZYtech). Then, we diluted the 1:10 the cDNA and carried out a qPCR using Power-up SYBR green (ThermoFisher) and the following primers: BMPR2 (forward 5’-AGAGACCCAAGTTCCCAGAAGC-3’, reverse 5’-CCTTTCCTCAGCACACTGTGCA), HDAC2 (forward 5’-CTCATGCACCTGGTGTCCAGAT-3’, reverse 5’-GCTATCCGCTTGTCTGATGCTC-3’), ALAS1 (forward 5’-AGTGTGAAAACCGATGGAGG-3’, reverse 5’-CGATCATACTGAAAAGTGGAAACAG-3’), YWHAZ (forward 5’-ATGCAACCAACACATCCTATC-3’, reverse 5’-GCATTATTAGCGTGCTGTCTT-3’). We normalized the expression using the ddCT method with *YWHAZ* and *ALAS1* as reference genes.

We plated HeLa cells in a 6-well plate. After achieving an 80-90% confluence, we transfected them with a 40 µM concentration of miR-3168, scramble or miR-3168 + ago-miR-3168 (IDT, USA). After 48 hours, we extracted the protein using RIPA buffer and quantified it using Pierce BCA protein Kit (ThermoFisher).

We prepared 20 µg of protein in Laemlii’s sample buffer (BioRad) containing 5% B-mercaptoethanol (Sigma) and heated it at 95°C for 5 minutes. Proteins were then separated by SDS-Page using a 7.5% mini-Protean TGX precast gel (BioRad). After electrophoresis, we transferred the proteins into a PVDF membrane using a Trans-Blot Turbo Transfer pack (BioRad) in a Trans-Blot Turbo system (BioRad) for 7 minutes at 1.3 A. We then incubated the membrane for 1 hour in a blocking buffer composed of 5 % w/v nonfat milk in Tris-buffered saline (TBS) containing 0.2 % Tween20 (TBS-T). Immunoblotting was carried out by incubating the membrane with a 1:1000 dilution of anti-BMPR2 antibody (Abcam #ab130206) for 1 hour at RT. Then, we washed the membrane three times in blocking buffer and incubated it with a 1:10000 dilution of anti-Mouse IgG-HRP antibody (Abcam, #205719). Finally, we washed the membrane with TBS and developed the blot using Clarity Western ECL substrate (BioRad). We stained the membrane with Coomassie Brilliant Blue and used total protein as loading control. Imaging was carried out in a ChemiDoc (BioRad) digital camera-based imaging system.

### Flow cytometry

We transfected PAECs in passages 3-5 with: miR-3168, scramble or miR-3168 + ago-miR-3168a at 40 µM concentration. After 24h, we changed the media for EGM supplemented with 2% FBS for 24h. Then, we harvested the cells and the media, pelleted them, and washed them twice in PBS without Calcium. Finally, we resuspended the pellets with Annexin binding buffer containing an anti-Annexin-FITC and 7-AAD (FITC Annexin V Apoptosis Detection Kit with 7-AAD, BioLegend). We incubated the mixes for 15 min and run the cytometry using a Cytoflex (Beckman Coulter). Data analysis was performed using Cytexpert (Beckman Coulter).

### Tube formation analysis

After transfecting PAECs in the same conditions as before, we changed the media after 24h for EGM supplemented with 0.2% FBS. After 24h, we trypsinized the cells, counted them and plated in full growth media 20.000 cells per well in 96-well plates pre-coated with Matrigel (Sigma). We imaged the wells after 12 h using a Nikon TiU inverted microscope. Brightfield images were then analyzed using the Angiogenesis analyzer ImageJ plugin [25].

## Results

### Cohort description

The characteristics of the population is shown in Table 1. The table presents data on 110 patients diagnosed with various forms of PAH. Most of the patients were female (72%). The most common diagnosis was Idiopathic PAH (iPAH) at 83.6%, followed by familial PAH (FPAH) at 9.2%. Most patients were classified as Functional Class III (58.2%). The median time from diagnosis to the first visit was 7 days, with a range from 0 to 35 days. The mean 6-minute walk distance was 409 meters, and levels of NT-proBNP or BNP, as cardiac markers, were elevated. . In terms of hemodynamics, they also showed severe pulmonary hypertension and markedly elevated PVR indicating a serious condition.

**Table 1.**
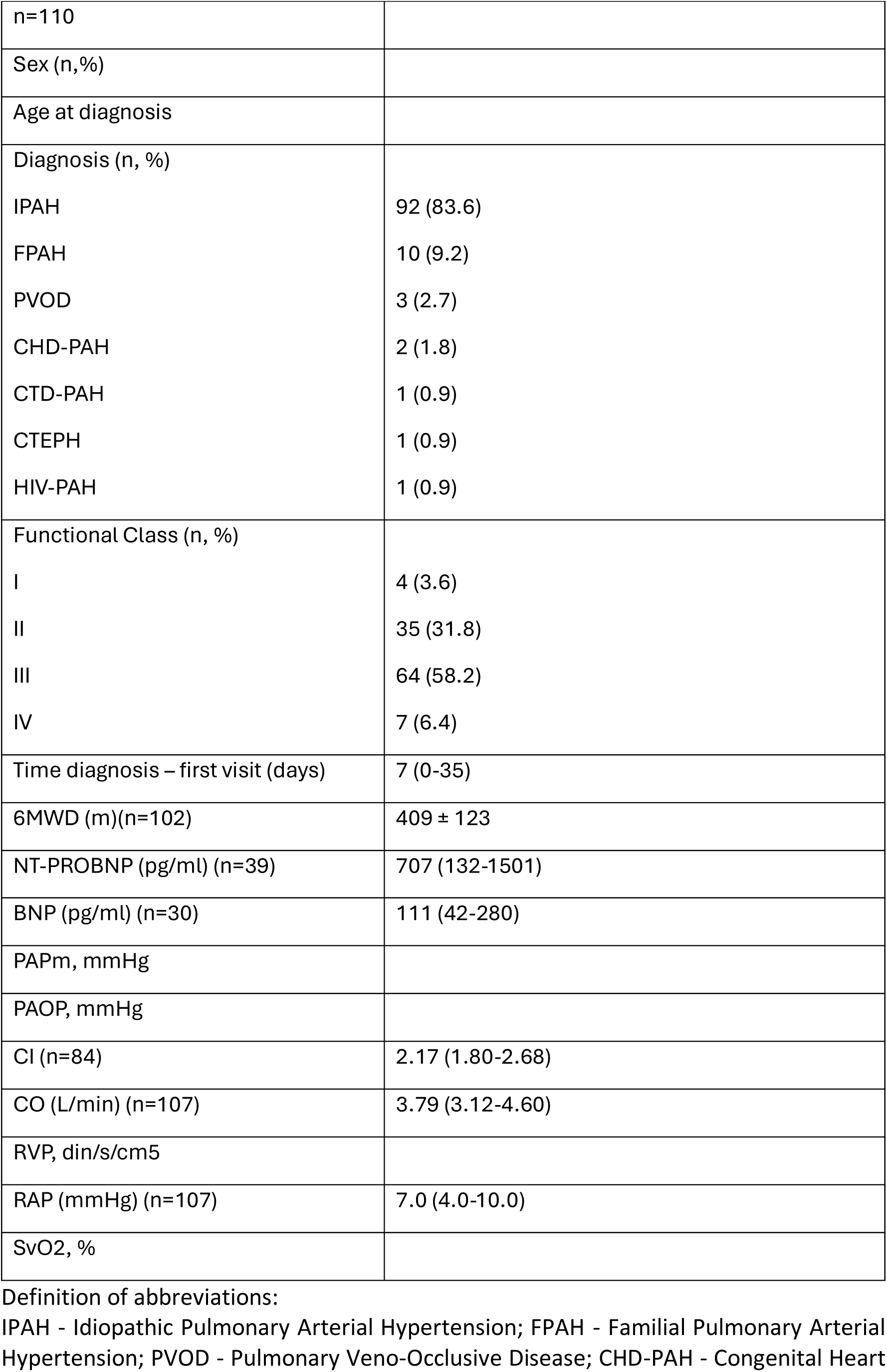
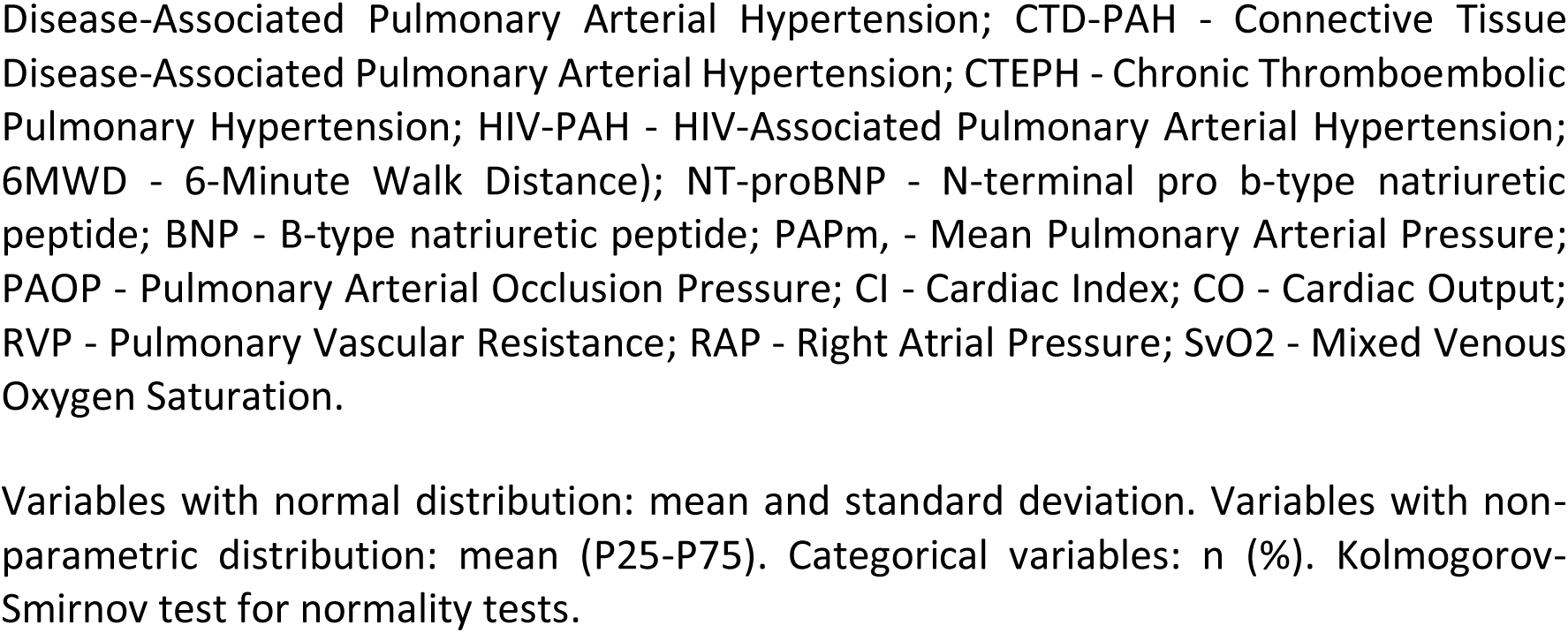
Characteristics of the population.

|  |  |
| --- | --- |
| n=110 |  |
| Sex (n,%) |  |
| Age at diagnosis |  |
| Diagnosis (n, %) |  |
| IPAH | 92 (83.6) |
| FPAH | 10 (9.2) |
| PVOD | 3 (2.7) |
| CHD-PAH | 2 (1.8) |
| CTD-PAH | 1 (0.9) |
| CTEPH | 1 (0.9) |
| HIV-PAH | 1 (0.9) |
| Functional Class (n, %) |  |
| I | 4 (3.6) |
| II | 35 (31.8) |
| III | 64 (58.2) |
| IV | 7 (6.4) |
| Time diagnosis – first visit (days) | 7 (0-35) |
| 6MWD (m)(n=102) | 409 ± 123 |
| NT-PROBNP (pg/ml) (n=39) | 707 (132-1501) |
| BNP (pg/ml) (n=30) | 111 (42-280) |
| PAPm, mmHg |  |
| PAOP, mmHg |  |
| CI (n=84) | 2.17 (1.80-2.68) |
| CO (L/min) (n=107) | 3.79 (3.12-4.60) |
| RVP, din/s/cm5 |  |
| RAP (mmHg) (n=107) | 7.0 (4.0-10.0) |
| SvO2, % |  |

Definition of abbreviations:
IPAH - Idiopathic Pulmonary Arterial Hypertension; FPAH - Familial Pulmonary Arterial Hypertension; PVOD - Pulmonary Veno-Occlusive Disease; CHD-PAH - Congenital Heart
Disease-Associated Pulmonary Arterial Hypertension; CTD-PAH - Connective Tissue Disease-Associated Pulmonary Arterial Hypertension; CTEPH - Chronic Thromboembolic Pulmonary Hypertension; HIV-PAH - HIV-Associated Pulmonary Arterial Hypertension; 6MWD - 6-Minute Walk Distance); NT-proBNP - N-terminal pro b-type natriuretic peptide; BNP - B-type natriuretic peptide; PAPm, - Mean Pulmonary Arterial Pressure; PAOP - Pulmonary Arterial Occlusion Pressure; CI - Cardiac Index; CO - Cardiac Output; RVP - Pulmonary Vascular Resistance; RAP - Right Atrial Pressure; SvO2 - Mixed Venous Oxygen Saturation.
Variables with normal distribution: mean and standard deviation. Variables with non-parametric distribution: mean (P25-P75). Categorical variables: n (%). Kolmogorov-Smirnov test for normality tests.

### Differential Expression analysis

We detected 29 different miRNAs that were statistically significant (FDR > 1%) and had more than a ± 50% difference between patients and controls using DESeq2 (Figure 1C), principal component analysis showed PAH patients clustering together and a lot of variability between the donors (Figure 1B). We then run again the DE analysis using EdgeR, merged both datasets and compared the DE miRNAs. We found a match for 26 of them woth consisntent FC and minimal variation, which still exceeded the FDR (Supp. Figure 1).

Based on FDR and FC, we selected 13 candidate miRNAs (let-7a-5p, let-7b-5p, let-7c-5p, let-7f-5p, miR-9-5p, miR-31-5p, miR-3168, miR-203a-3p, miR-1468, miR-517b-3p, miR-6735, miR-657, miR-6853-3p) for a prevalidation in 50 patients and 50 controls. To normalize the expression of the different miRNAs, we used the DE data to look for miRNAs that had very little difference between IPAH and controls and were highly abundant. This way we selected miR-16a and miR-103 as reference miRNAs.

### Biomarker Prevalidation

#### Let-7a and Let-7f are downregulated in IPAH

Out of the 13 candidate miRNAs, only 7 amplified correctly by qPCR. From the downregulated miRNAs, only Let-7a and Let-7f kept showing lower levels than the controls and were statistically significant (Figures 2A-G).

**Figure 2.**
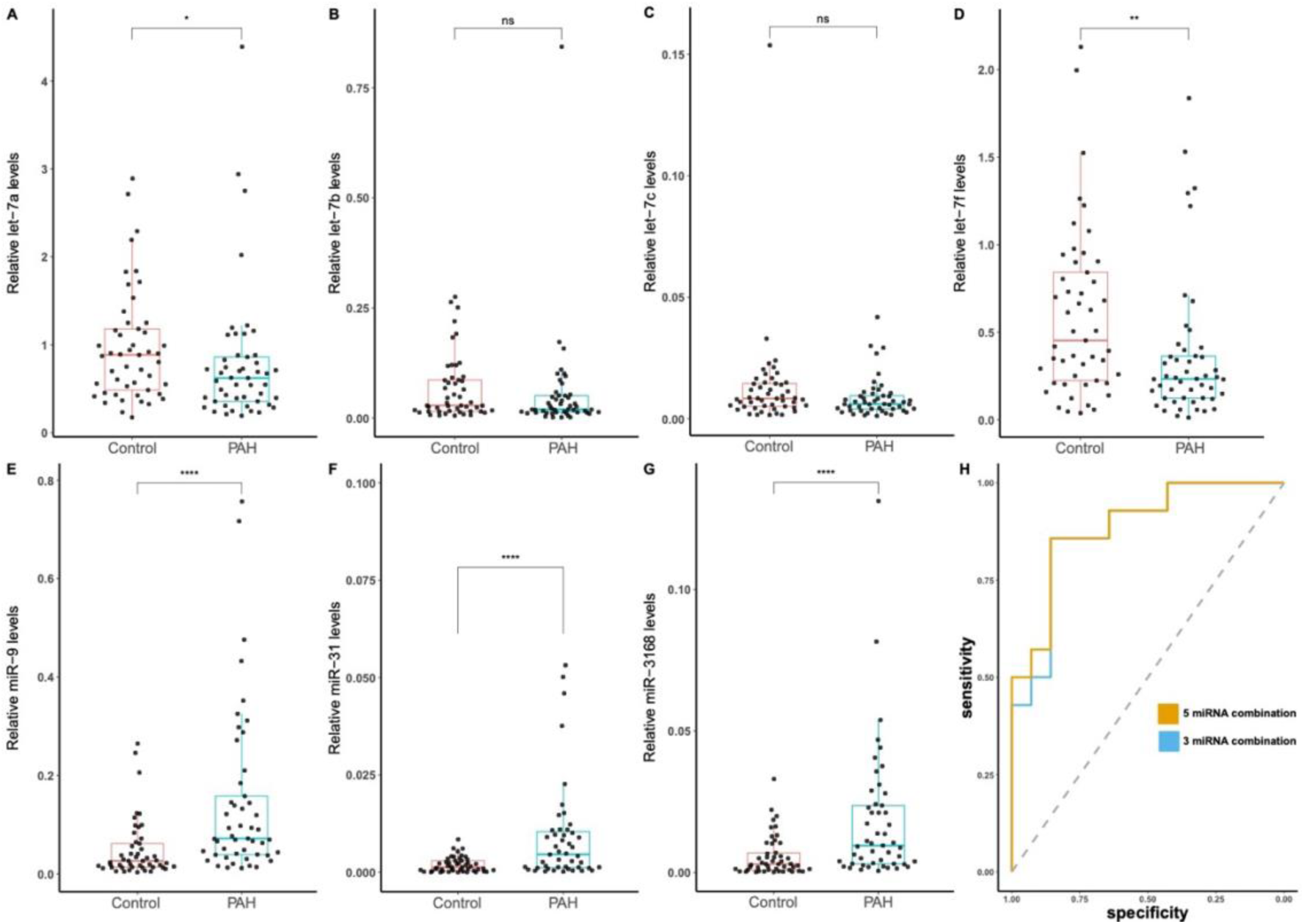
Prevalidation of 13 miRNAs showed detectable levels for only 7 of the candidates, A, B, C, D) Comparison of the let-7 family member’s levels in the prevalidation cohort. Let-7a and Let7f show statistically significant differences between the groups. E, F, G) Comparison of the significantly upregulated miRNAs detected in the sequencing (miR-9, miR-31, miR-3168) levels in the prevalidation cohort. H) ROC curve analysis and AUC of the two models evaluating PAH diagnosis by logistic regression of a combination of 5 miRNAs (orange; miR-9, miR-31, miR-3168, let-7a and let-7f) or 3 (blue; miR-9, miR-31 and let-7f). Data are shown as box plots representing median ± quartiles. Dots represent individual values for each patient/control. *P* values determined by a unpaired t-test (ns = not significant, * p > 0.05, ** p > 0.01, **** p > 0.0001).

We carried out an *in silico* binding prediction using miRbase [24]. The results showed that the whole Let-7 family targeted *EDN1*.

#### MiR-9, miR-31 and miR-3168 are upregulated in IPAH

The levels of miR-9-5p, miR-31-5p and miR-3168 were higher in PAH patients when compared with the controls, the differences were statistically significant for all of them (Figures 1E-G). MiR-9-5p has been associated with cell proliferation and apoptosis resistance in different types of cancers. It is predicted to bind to *ATP13A3*, one of the newest PAH genes.

#### Classification models

We used a lasso regression to analyze which candidate miRNA contributed the most to diagnostic efficiency in the prevalidation cohort (Table 2).

**Table 2.** Lasso regression miRNA priorization.

| miRNA | Score |
| --- | --- |
| miR-9-5p | 0.45814792 |
| let-7a-5p | -0.40188783 |
| miR-31-5p | 0.16811012 |
| let-7f-5p | -0.09602841 |
| miR-3168 | 0.02555715 |
| let-7b-5p | 0 |
| let-7c-5p | 0 |

Using the predictions from this model we calculated the area under the curve using a ROC curve (Figure 2H). Overall, this model showed a cut point with high sensitivity/specificity and AUC of 0.862. We repeated the model using a logistic regression and the results improved slightly. We obtained an AUC of 0.888 with the same 5 miRNAs (Figure 2H, orange line). Then, we proceeded to delete from the model the other miRNAs to simplify the miRNA signature panel. By using three (Let-7a, miR-9 and miR-31) the model had an AUC of 0.878 (Figure 2H, blue line). The simplified version of the miRNA panel was almost identical to the full version, so we opted for testing only these three miRNAs in the rest of the cohort.

### Biomarker validation

We analyzed the expression of the three prioritized candidate miRNAs in 60 more patients and 70 controls. After normalizing and mixing the data from the complete validation cohort, miR-let-7a loss significance. While miR-9 and miR-31 are overexpressed in the full cohort (boxplot Figure 3A-C).

**Figure 3.**
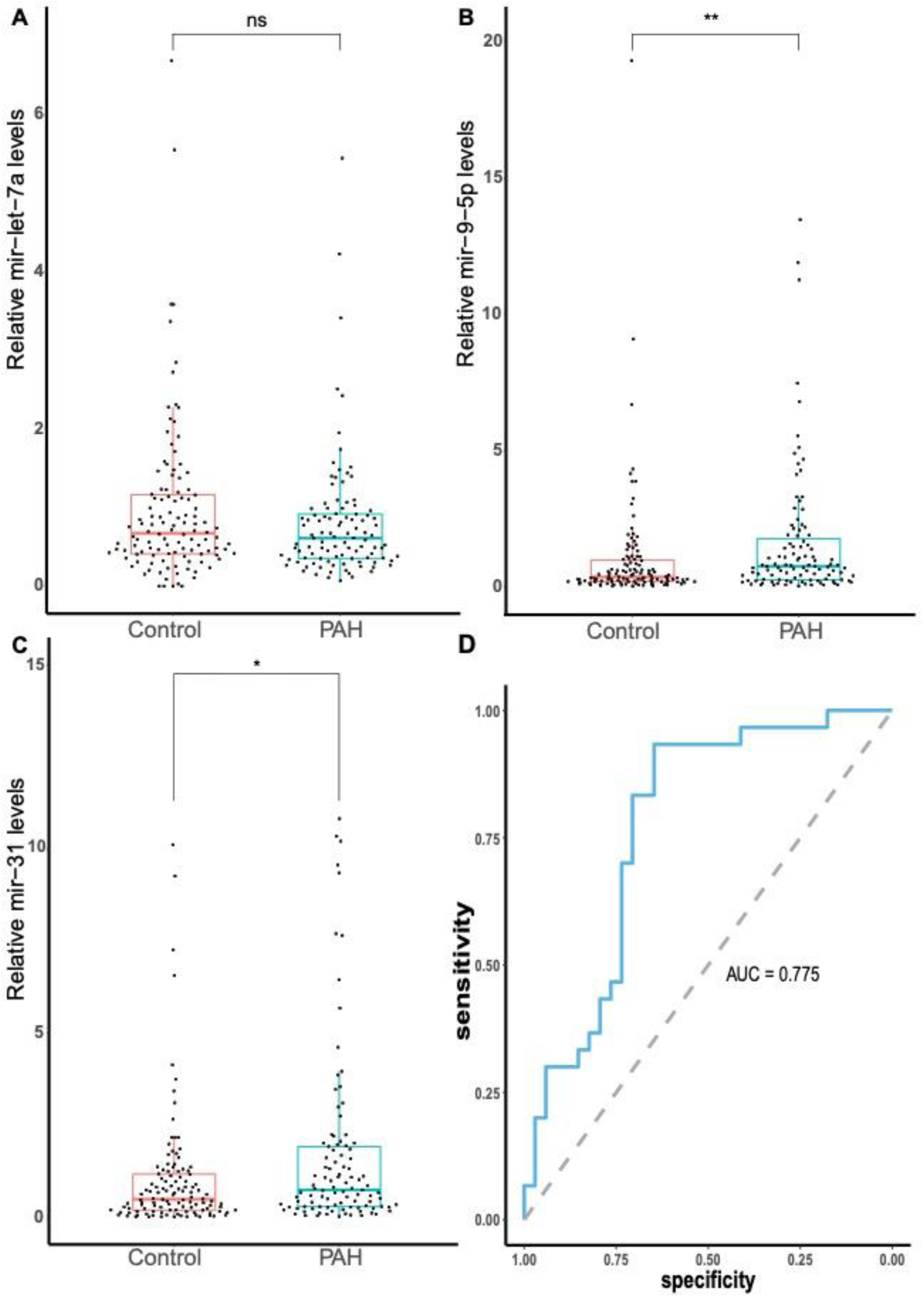
Diagnostic performance of the 3 miRNAs model in the full cohort (n=230). A) Quantification of miR-let-7a levels. B) Quantification of miR-9-5p levels. C) Quantification of miR-31 levels. D) ROC curve analysis and AUC of the 3 miRNAs model evaluating PAH diagnosis by logistic regression (miR-9, miR-31 and let-7f). Data were normalized using miR-16a and miR-103a levels, the box plot s how median ± interquartile ranges. Dots represent individual biological replicates. *P* values determined by a unpaired t-test (ns = not significant, * p > 0.05, ** p > 0.01).

#### Panel validation

We used the model generated during the prevalidation and applied it to the validation set. As expected, after plotting the levels of the miRNAs, the classification worsened obtaining an AUC of 0.736 with deteriorated cut points (Figure 3D).

### Risk analysis and correlation with miRNA levels

To analyze the possibility of using these miRNA panel to predict prognosis, we calculated the Risk following the method described by COMPERA, as well as plotting the miRNA levels depending on the functional class at the moment of blood draw. Let-7a was the only miRNA whose levels changed with functional class, showing an increase in groups II and III (Supp. Figure 2A-C). There was no strong correlation between any of these miRNAs and the Risk (Supp Figure 2D and 3). Furthermore, the analysis of miRNA levels by PAH subtype did not show any difference (Supp. Figure 4).

### Target prediction and functional analysis of miR-3168

We looked for targets with scores >80. We looked for PAH related genes, we only found *EDN1* in the let-7 cluster, *ATP13A3* for miR-31-5p and *BMPR2* for miR-3168. Considering that miR-3168 was unknown in the literature; we decided to test if its effects were real. We choose two of the predicted targets of miR-3168 (*BMPR2* and *HDAC2*) and checked their mRNA levels after transfecting different amounts of a miR-3168 mimic or a scrambled miRNA. The results showed that at a 20 µM concentration, both targets showed decreased mRNA levels (Supp. Figure 5). *BMPR2* showed relative mean levels of 0.745 ± 0.167 at 20 µM and 0.406 ± 0.135 at 40 µM. In the case of *HDAC*, it showed mean levels of 0.579 ± 0.11 at 20 µM and 0.589 ± 0.27 at 40 µM.

To confirm the relationship, we selected the 40 µM concentration to test the BMPR2 protein levels after transfecting in the same conditions, but in this case, we added agomiR-3168 as an additional control. We found statistically significant differences between miR-3168 treatment showed lower relative levels (0.25 ± 0.11) when compared against the scramble (1 ± 0.22). At a protein level, miR-3168 inhibits BMPR2 protein levels up to a 75 % (Figure 4A). When compared with the scramble, miR-3168 treatment showed lower BMPR2 levels (0.25 ± 0.1, adj. p > 0.05) while the treatment of Ago-miR-3168 + miR- 3168 showed levels very similar to the scramble (1.24 ± 0.47).

**Figure 4.**
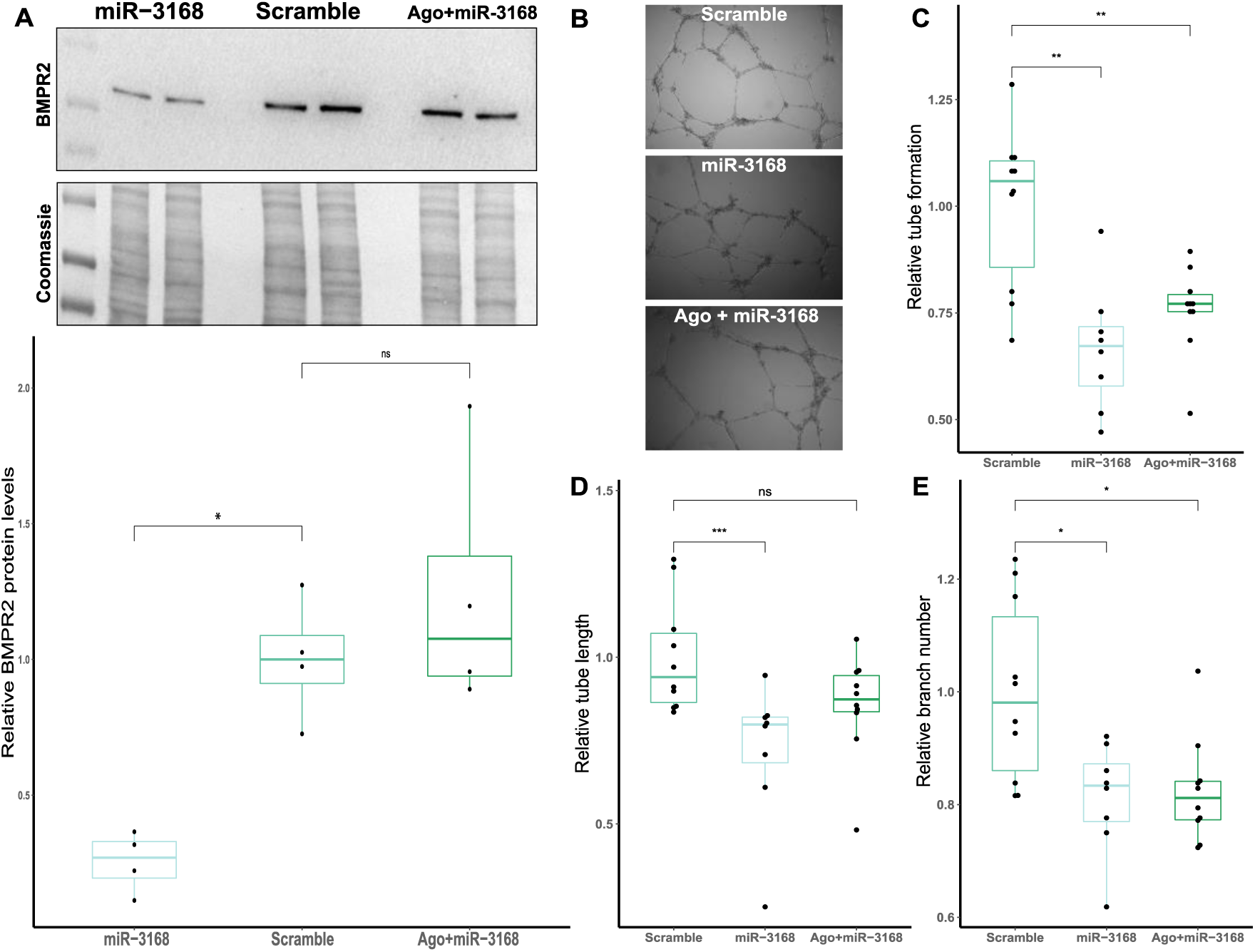
miR-3168 reduces BMPR2 expression and impairs angiogenesis. A) Box plot representing the quantification of BMPR2 protein levels after treatment with miR-3168, a scramble miRNA and Ago-miR-3168 + miR-3168 (n = 4). A representative western blot for BMPR2 and its loading control using Coomassie staining are depicted. B) Effect of miR-3168 on angiogenesis evaluated by tube formation analysis (n = 10). Representative image of the tube formation assay. C) Quantification of the relative number of tubes formed by treatment. D) Quantification of the relative length of the tubes. E) Quantification of the relative number of branches sprouted. Data were relativized against the scramble, the box plot s how median ± interquartile ranges. Dots represent individual biological replicates. *P* values determined by a one-way ANOVA (ns = not significant, * p > 0.05, ** p > 0.01, *** p > 0.001)

### Cell viability and apoptosis assay in HUVECs

We found no differences in cell viability and apoptosis between the treatments and the control (n = 7; Supp. Figure 6).

### Tube formation analysis

Treatment with miR-3168 reduced tube formation by a 33.8% when compared with the WT (Figure 4B, C, p > 0.01); adding AgomiR-3168 increased slightly tube formation (24.2 % reduction, p > 0.01). In terms of tube length (Figure 4D), miR-3168 decreased tube length a 28% (p >0.001) while the addition of the agonist reduced tube length loss to 14.5%, showing very similar results to WT. Regarding branching, miR-3168 treatment reduced about a 15% the number of branching sites (Figure 4E, p >0.05) and treatment with the agonist did not prevent it.

## Discussion

Early diagnosis of PAH remains a significant challenge. In most cases, patients are diagnosed when the pulmonary vasculature is about to collapse. We have known for several years that the sooner the treatments start, the better the prognosis. We are not only speaking of life expectancy, but also quality of life. That’s why we urgently need diagnostic tools that can be used before patients exhibit symptoms. Furthermore, the diagnosis of PAH must always be confirmed through right heart catheterization, an invasive procedure that, like any intervention, carries inherent risks.

In this article we aimed to characterize the circulating miRNome of IPAH patients in early functional classes, discover novel miRNAs that could be involved in the pathogenesis and develop an efficient diagnostic signature.

We were able to identify 26 miRNAs that were DE between IPAH and controls. Then, from these 26 we limited to 7 the number of candidates for the classification models and used *in silico* predictions to discover interesting target genes.

In our DE analysis we detected the let-7 family members’ let-7a, let-7b, let-7c and let-7f to be downregulated in the patients. This was not the first time that members of this family were related to PAH, as let-7a had been reported to be downregulated under hypoxia and let-7f after monocrotaline treatment in a rat model [26], while the closely related let-7i-5p was upregulated in IPAH patients [27]. From this cluster, let-7b and let-7c did not show statistically significant downregulation in our prevalidation cohort, while let-7a and let-7f did so. However, in the validation set let-7a also lost its significance. The miRNAs of the let-7 family are known repressors of cell cycle progression and proliferation [28]. They are well characterized tumor repressors, and their downregulation is associated with worse prognosis in different types of cancers [29–31]. If we couple this data with the fact that let-7 alter the levels of ET-1 by binding to the 3’UTR of *EDN1* [32,33], we can hypothesize that a downregulation of this miRNAs could play a role in the development of PAH. An increase of vasoconstriction via ET-1 production and increased PAEC proliferation due to altered cell cycle would fit very well with PAH development.

In the case of the upregulated miRNAs, we had many more candidates, but the number of copies we detected in the sequencing was very variable, with some individuals having thousands of copies and others none. Thus, we were only able to amplify miR-9-5p, miR-31-5p and miR-3168. All of them were overexpressed in PAH patients in the prevalidation cohort, and the prioritized miRNAs (miR-9 and miR-31) were also overexpressed in the full cohort, although the differences in the validation cohort alone were lower.

The overexpression of miR-9 has been reported in different cancer types. It has been well characterized in glioma, where miR-9 is described to increase proliferation, migration and invasion of the cancer cells [34]. They also secrete miR-9 the exosomes to increase angiogenesis, something that was proved *in vitro* by the increase of HUVEC proliferation and tube formation after treating them with glioma-derived exosomes [34]. This promotion of angiogenesis happens due to miR-9 inhibition of S1P_1_ [35]. But not only endothelial cells can be influenced by this miRNA, fibroblasts can be involved in a crosstalk with tumors, increasing its motility and enhancing epithelial-mesenchymal transition [36,37]. Altogether, miR-9 could be promoting PAEC proliferation and helping in the development of PAH.

High levels of miR-31-5p have been associated with tumor progression in several types of cancers [38–40]. There are several circular RNAs that act as a sponge for this miRNA, when their levels are low the prognosis is worse [41,42]. The pathways regulated by miR-31-5p are diverse, but *in vitro* experiments have shown that it enhances proliferation, migration and inhibited apoptosis [41]. But miR-31-5p has not only been reported in cancer, in aortic dissection/aneurism miR-31-5p expression downregulated the expression of contractile genes by lowering myocardin levels [43]. Although, miR-31-5p is also supposed to lower inflammation by downregulating Protein Kinase C ε and indirectly the NF-κB pathway [44]. Recently, miR-31-5p was reported to be upregulated in the right ventricle of PAH patients, especially in male patients, where its expression was regulated by a network of circular RNAs and lncRNAs [45]. Besides, miR-31-5p could target *ATP13A3*, whose haploinsufficiency causes PAH [46,47]. Thus, the implication of this miRNA in PAH pathogenesis make sense if we focus in proliferation/inhibition and how it is reported to interact with vascular smooth muscle cells [41,43], it could help in the overall proliferative environment of PAH-development with enhanced SMC/PAEC/AF proliferation and migration. However, the inhibition of inflammatory responses may be protective effect in this case.

MiR-3168 is the less known miRNA that we studied, some of the targets predicted for it were interesting, as several BMP receptors were involved (*BMPR2* and *BMPR1A*). When we analyzed the *BMPR2* mRNA, we found that it decreases after treating the cells with miR-3168, the same happened with *HDAC2* levels (another of the miRDB-predicted targets). The protein levels of BMPR2 were also downregulated. Furthermore, *BMPR2* inhibition is one of the better understood causes of endothelial dysfunction [48–50]. Nevertheless, HDAC2 is upregulated in pulmonary adventitial fibroblasts from IPAH patients and it is mostly expressed in SMCs [51]. Our functional studies of miR-3168 pointed out that it alters angiogenesis *in vitro*, which supports our finding of it targeting *BMPR2*, but the relationship between this miRNA and PAH should be further studied.

Our miRNA extraction discarded big exosomes and apoptotic bodies with the first centrifugation, but we have extracted miRNAs from smaller vesicles. Proof of this is that a big part of let-7a is linked to vesicles [52], so we cannot be sure whether or not this miRNA dysregulation is showing crosstalk between PAH-specific cellular types.

The study design had limitations, during the miRNA discovery step we could not get patients as homogenous as we would have liked. Although we matched ages and sex, it was not sufficient to obtain better results. Another point to consider was treatment, as our patients were not naïve, we tried to address this issue by using only patients in functional NYHA classes I and II in the discovery cohort.

## Conclusions

In summary, we identified differentially expressed miRNAs between patients and controls and successfully validated these findings within our cohort. Our results highlight the downregulation of the let-7 family in PAH patients and the overexpression of miRNAs related with proliferation (miR-9-5p and miR-31-5p). Additionally, we found that miR- 3168 is upregulated in PAH patients, targets *BMPR2*, and impair angiogenesis *in vitro*. Therefore, it could play a role in the development of PAH.

## Data Availability

All data generated in this article is available upon request. RNA-seq derived data is freely available at GSE222022, count matrix and DESeq2 analysis tables can be accessed in the supplementary dataset.

https://www.ncbi.nlm.nih.gov/geo/query/acc.cgi?acc=GSE222022

## Declarations

### Ethics approval and consent to participate

The study was conducted after the approval of the *Comité de ética da Investigación de Galicia* (CEIC Galicia).

### Contributions

MLD and DV designed and conceptualized the study. MLD performed all the experimental and analysis work. AIG assisted in the validation cohort analyses. MLD wrote the manuscript. AB, CV, JAB and IB collected samples collection and retrieved clinical history. DV supervised the study. All authors approved the manuscript.

## Acknowledgements

We thank all the patients that participated in this study and all the clinical personnel implied in sample retrieval and the IdiBaps Biobank. We thank the REHAP investigators for their effort to recruit and manage the patients. We thank Fundación Contra la Hipertensión Pulmonar for their continues support and effort. We thank Laura Muinelo Romay for providing us with the controls for the discovery cohort. We thank Merce Peleteiro from the Cytometry core facility of CINBIO for her technical help. In loving memory of Guillermo Pousada.

