## Supplementary material for "Overexpression of miR-3168 impairs angiogenesis in Pulmonary Arterial Hypertension: Insights from circulating miRNA analysis": Supp. Figure

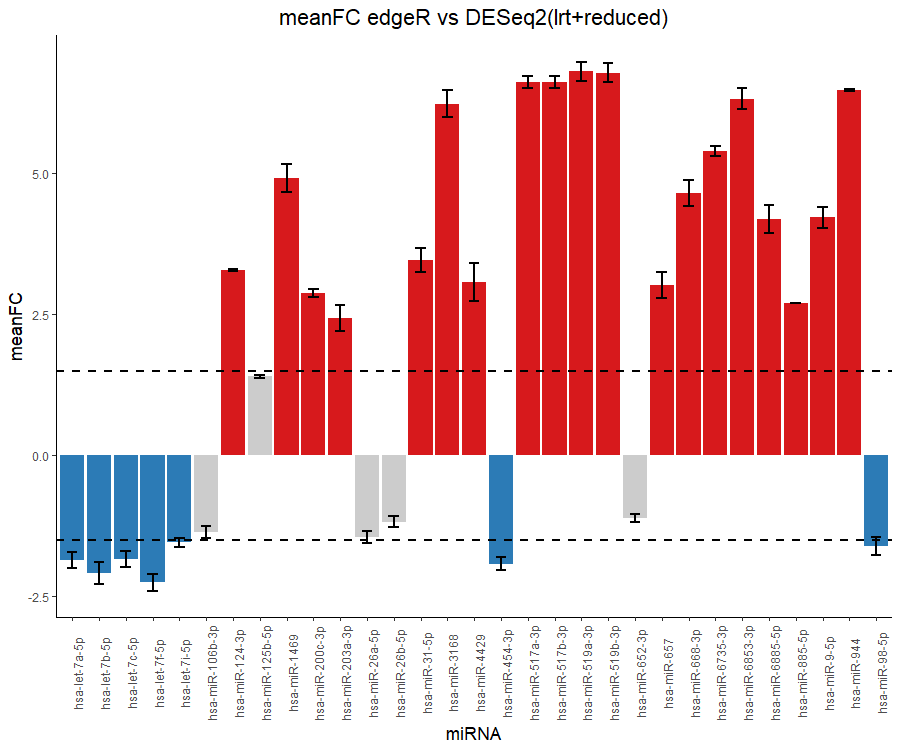

Supp. Figure 1. Comparison between DESeq2 and EdgeR DE results. Data are mean ± standard deviation. Both methods selected the same differentially expressed targets with minimum variability.

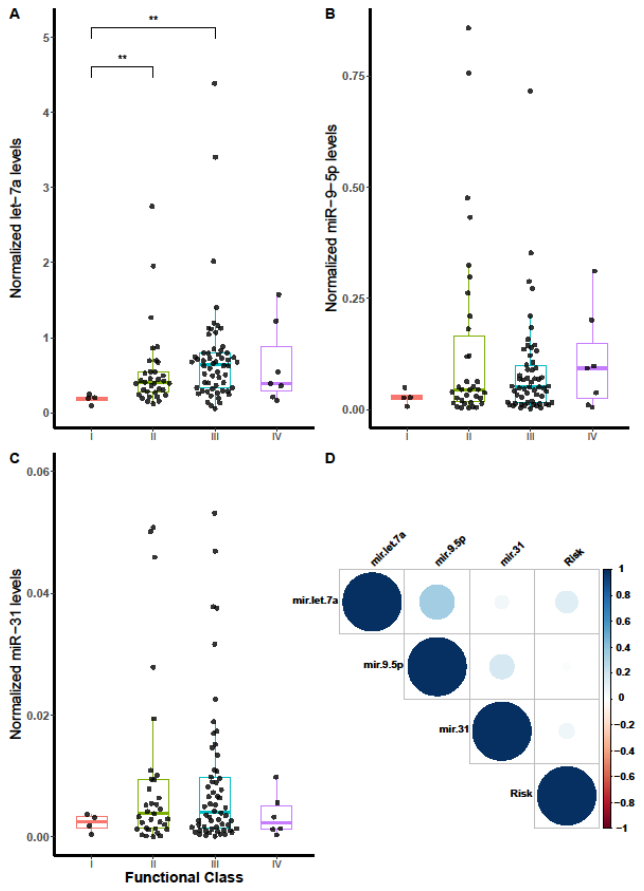

Supp. Figure 2. miRNA expression by functional class and correlation with COMPERA Risk. Levels of let-7a (A), miR-9-5p (B) and miR-31 (c) by functional class. D) Correlation of the levels of the miRNAs used in the panel with the risk according to COMPERA, only miR-9-5p showed moderate correlation. Data are shown as box plot depicting media ± interquartile ranges, individual samples are shown as dots.

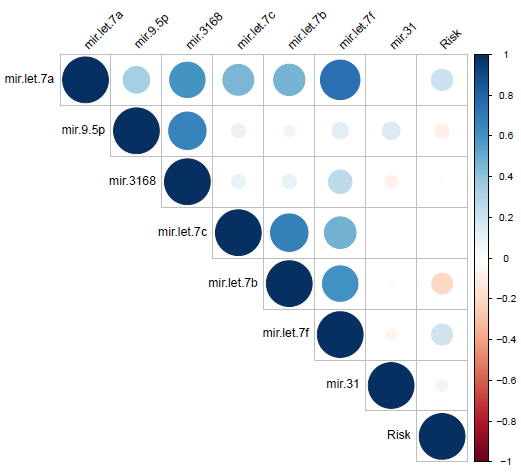

Supp. Figure 3. Correlation analysis of all the miRNAs analyzed and Risk.

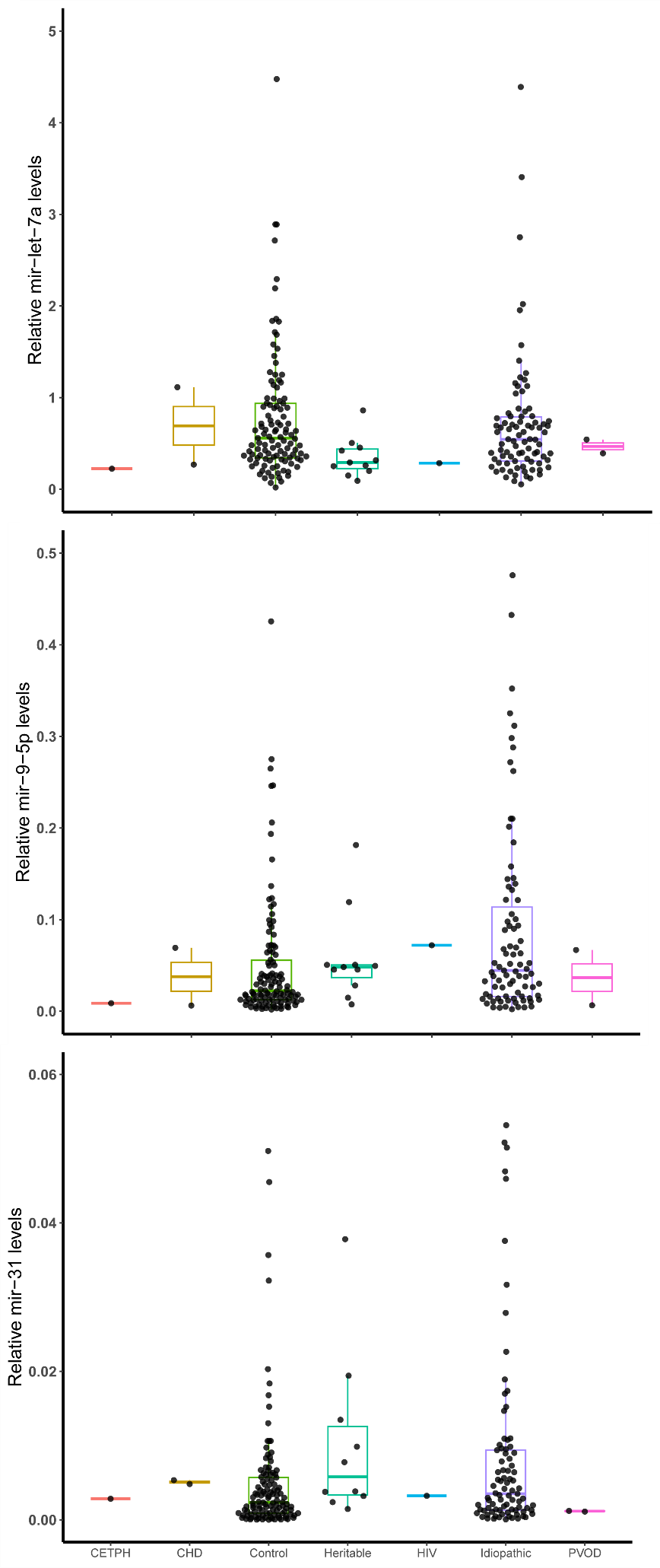

Supp. Figure 4. MiRNA levels by PAH etiology. A) Relative miR-let7a levels. B) Relative miR-9-5p levels. C) Relative miR-31 levels. Data are shown as box plot depicting media ± interquartile ranges, individual samples are shown as dots.

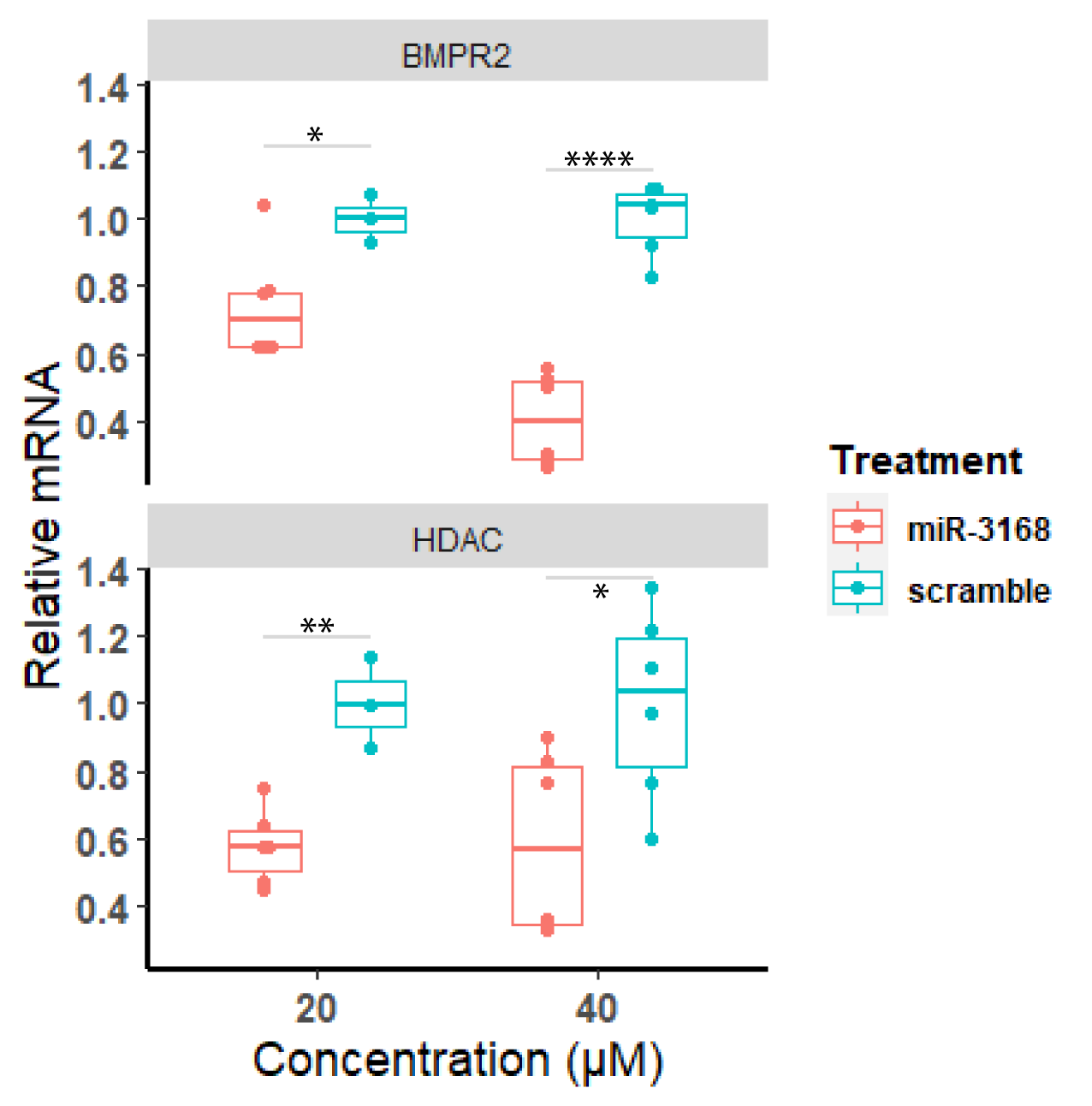

Supp. Figure 5. Analysis of the mRNA levels of miR-3168 targets after the addition of this miRNA in different concentrations (n = 6). Data were relativized against the scramble with the same concentration, data are shown as box plot depicting media ± interquartile ranges, and individual qPCR replicates are depicted as dots. p > 0.05 *, p > 0.01 **, p > 0.0001 ****.

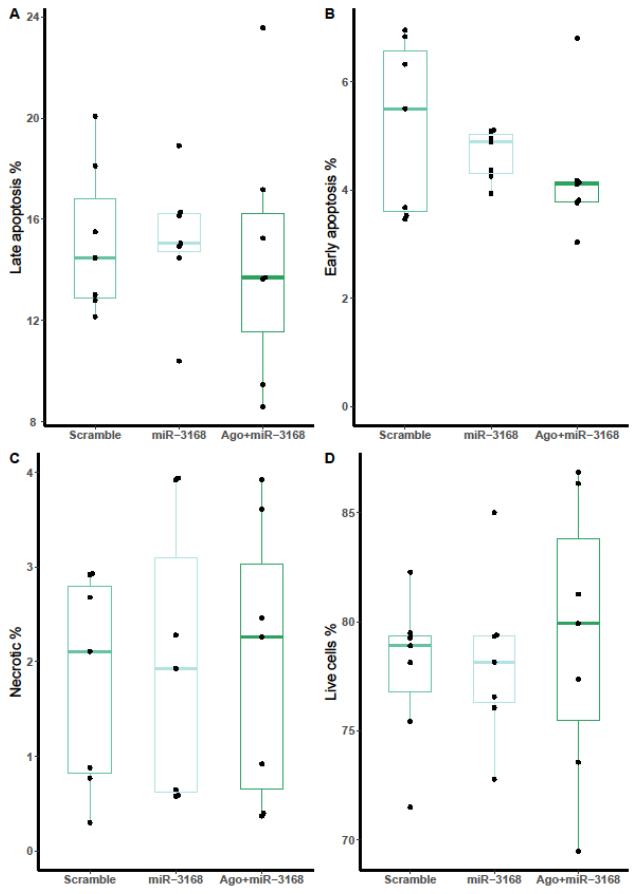

Supp. Figure 6. Flow cytometry analysis of cell viability. The Anexin V analysis showed no significant differences in the amount of late apoptotic, early apoptotic, necrotic and live cells between treatments. Data are shown as box plot depicting media ± interquartile ranges, individual replicates are shown as dots (n = 8).
